# A u-shape association of serum uric acid with gout in US adults

**DOI:** 10.1101/2022.07.12.22277537

**Authors:** Yan Xinmiao, Sun Taolan, Zhao Zixuan, Luo Yanrong, Zhao Hanbing, Zhao Han, Li Miaojing

**Author notes:** Corresponding author (LMJ).

## Abstract

**Background:** Serum uric acid (SUA) level is the strongest determinant of gout, and the relationship between hyperuricemia and gout has been found. However, the association between the across-full range of SUA level and gout also remains uncertain. We aimed to investigate the trend across an across-full range of SUA level and gout in the general population.

**Materials and Methods:** This study included 4,738 US adults from The National Health and Nutrition Examination Survey (NHANES). A generalized additive model was used to fit the non-linear trend between SUA level and gout.

**Results:** 2,209 males and 2,469 females were included in our study, with the age range of 20∼80 years. The range of SUA level was from 0.8 to 15.1 mg/dL. About 5.9% of participants had gout. There was a U-shape association between SUA level and gout in univariable GAM (EDF = 3.881, p <0.001) and multivariable GAM (EDF = 3.795, p <0.001). The lowest risk of gout was observed in participants with SUA level of 5-6mg/dL. We evaluated the trend of SUA level with gout by gender. Both males and females with high SUA level have a high prevalence of gout. Male have a high prevalence of gout with low SUA level. The SUA level and prevalence of gout support a U-shape in males, while females show a linear shape.

**Conclusion:** This study showed that low as well as high SUA are associated with gout. Hypouricemia is a candidate predictor of gout in US adults.

## Introduction

According to the Global Burden of Disease (GBD) 2017, there were 41.2 million (95% *UI:* 36.7 million∼46.1 million) adults with gout worldwide. In 2017, the age standardized point prevalence estimate and the increase of age-standardized point prevalence estimate of gout in the United States ranked among the top three in the world[1]. A prospective cohort study showed that the all-cause standardized mortality in gout patients was 2.21 (95% *CI:* 1.68-2.74)[2]. In addition, gout is associated with several diseases, including joint pain, chronic kidney disease (CKD), hypertension, type 2 diabetes, dyslipidemia, etc[3], which has effect on the life [4], productivity[5, 6] and health-related quality of life (HRQOL)[7, 8]. High uric acid (hyperuricemia) is the strongest determinant of gout, and many studies have proved the existence of this association [9-11]. SUA level can be maintained at a certain target level through urate-lowering treatment (ULT), effectively reducing the incidence of gout[12, 13]. Although some research on high SUA level and gout has been reported, the underlying association between across-full range of SUA level and gout is still uncertain. There is currently a lack of consensus regarding the optimal range for SUA levels and few studies have evaluated the prevalence of gout across the full range of SUA level in the US adults.

An analysis of the time of first gout occurrences using Cox proportional hazards model in a cohort study in Japan showed that the HR of asymptomatic hyperuricemia (5.0 < SUA ≤ 6.0 mg/dL) of ULT was the lowest, at 0.45 (95% *CI:* 0.27-0.76), compared with untreated subjects (SUA ≥ 8.0 mg/dL). In subjects with asymptomatic hyperuricemia, controlling SUA can reduce or eliminate gout occurrences[14]. In another Mendelian randomization study, it presented convincing evidence that the increased risk of gout was associated with high SUA level (*n*=71 501, *OR*: 5.84; 95% *CI*: 4.56-7.49, *p*<0.01)[15]. Meanwhile, studies have shown that before the age of 65 years, male have a fourfold greater incidence of gout than female [10, 16-18].However, the risk of gout development in patients with hypouricemia has not yet been clarified. D’silva et al reported that the risk of all-cause mortality risk among US males with SUA level <4 mg/dL was about 30% higher[19]. This indicates that it is necessary to further study the relationship between SUA level and gout and its impact on males and females.

Therefore, based on the National Health and Nutrition Examination Survey (NHANES), we explore the trend between SUA level and gout.

## Materials and methods

### Data Source and Study Population

The National Health and Nutrition Examination Survey (NHANES) is a unique source of national data on the health and nutritional status of the US population, collecting data through interviews, standard exams, and biospecimen collection. NHANES adopts a stratified and multi-stage sampling design, and the survey population involves each county in US. Each year, about 5,000 persons in 15 counties are selected to conduct a nationally representative sample survey. Informed consent was obtained from all participants.

NHANES 2017-2018 dataset was used in this study. There was a total of 9,254 participants being enrolled. Participants with age ≥20 years old were included for analysis. However, we excluded participants that had no uric acid data (*n*=3353), no gout data(*n*=987), missing in covariates variables (*n*=176). Eventually, 4,738 participants were included in our study and the distribution of the number of participants with different value of uric acid are shown in Fig 1.

**Fig 1.**
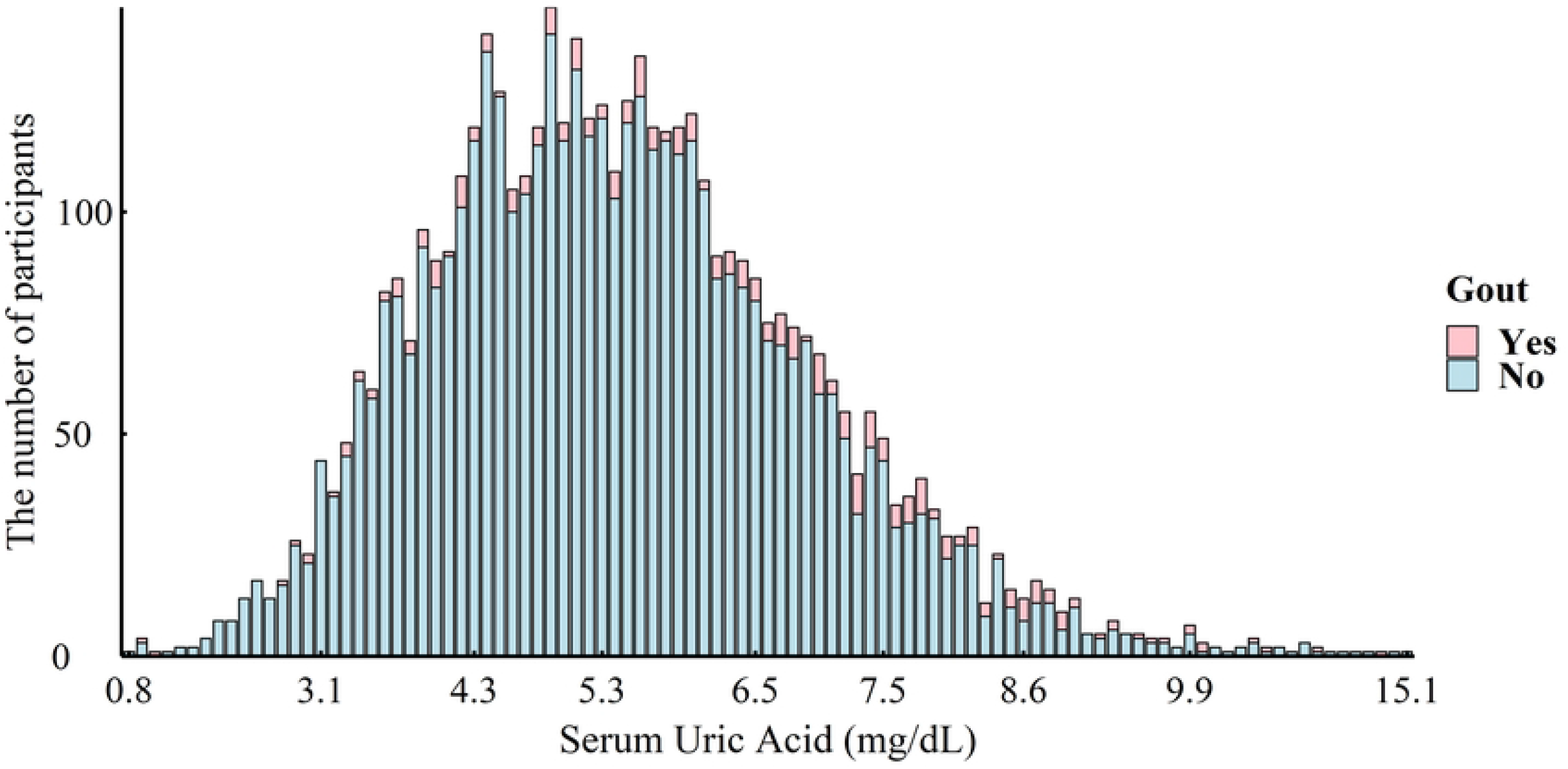
Distribution of the number of participants with different value of uric acid.

### Variable measurements

Socio-demographic factors and modifiable risk factor of gout were included in this study. We obtained the data of these variables from seven parts of following: “Demographic Data”, “Standard Biochemistry Profile”, “Alcohol Use”, “Blood Pressure & Cholesterol”, “Smoking - Cigarette Use”, “Kidney Conditions - Urology” and “Medical Conditions”.

### Gout

Gout was self-reported data. Whether participants have gout was defined using the question, “Has a doctor or other health professional ever told you that you had gout?”.

### Serum Uric acid level

In this study, we used the SUA level in the laboratory data as interested independent variable and recorded as a continuous variable. The detail process of laboratory data measurement was described in the NHANES database official website.

### Covariates

In this study, the covariates included gender, age, education, marriage, race, alcohol drinking, smoking, hypertension, diabetes and kidney disease. Age was divided into three groups: 20∼39, 40∼59, and 60 years or older. Education level was categorized into primary and secondary, high school, above high school. Marriage was grouped into four categories: married, widowed divorced or separated, never married, living with a partner. Race was divided into six categories: Mexican American, other Hispanic, non-Hispanic White, non-Hispanic Black, non-Hispanic Asian, other race - including Multi-Racial. Alcohol drinking was classified as never, almost every day, 3 to 4 times a week, 1 to 2 times a week, 1 to 3 times a month, and 1 to 11 times a year. Smoking was defined as “at least 100 cigarettes in a lifetime”. Diagnosis of hypertension, diabetes and kidney disease were determined with questions: “Have you ever been told by a doctor or health care provider that you have high blood pressure, diabetes or problems with high blood sugar, weak or failing kidneys”.

### Statistical Analysis

Descriptive statistics on participants’ characteristics were stratified by whether they had gout. Chi-squared test was utilized to determine the association between categorical variable. The generalized additive model (GAM) was used to assess the potential trend between SUA level and gout. The GAM approach is an extension of the generalized linear model (GLM), which is a generalized linear model with linear predictions involving sums of covariates (Wood, 2011). The smoothness of the model terms was estimated using penalized cubic regression splines for fitting. The model selection included the Akaike information criterion (AIC), the difference between EDF and ref. df, with lower values considered to be better-fitting models. As our response variable types were categorical variables, we chose “Logit” as our link function, and it was considered that the fixed effects were estimated by Restrictive maximum likelihood (REML). (http://doi.org/10.1080/01621459.2016.1180986)

Non-linear relationship of SUA level with gout was assessed through GAM. In our analysis, we fit three GAM models to NHANES data. The three models used gout (yes/no) as a binary response.

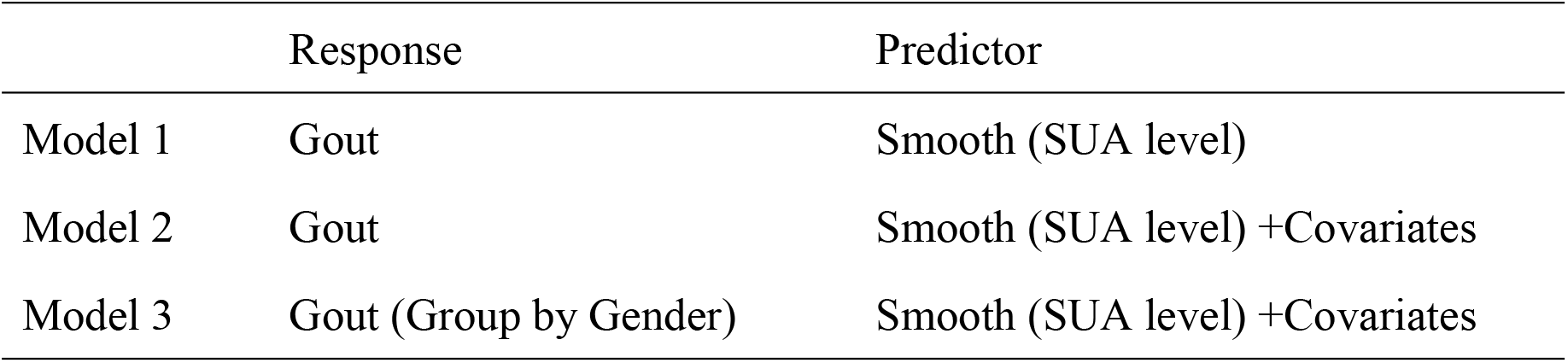

|  | Response | Predictor |
| --- | --- | --- |
| Model 1 | Gout | Smooth (SUA level) |
| Model 2 | Gout | Smooth (SUA level) +Covariates |
| Model 3 | Gout (Group by Gender) | Smooth (SUA level) +Covariates |

The reported values of effective degree of freedom (EDF) demonstrate the degree of curvature of the smooth. “EDF=1” means a linear pattern of the relationship. “EDF >1” is the sign of a more complex relationship between SUA level and gout. The Ref. df is close to the EDF when the GAM is reasonably fitted. The best explanation for the P-value of the smooth part is that a line parallel to the X-axis cannot pass through the 95% confidence interval of the curve.

SAS 9.4 (Statistical analysis system) was used for data cleaning and statistical description. GAM in our analysis was using the “mgcv” package (version 1.8–28) in R statistical software (version 4.1.1). The statistical tests were two-sided, and the significance level was set at 0.05.

## Results

### Characteristics of participants

A total of 4,738 eligible participants (2,209 males and 2,469 females) were included in our study, with the age range of 20∼80 years. About 48.12% were male, and 56.61% educated in above high school. A total of 50.65% of the sample were married. The range of SUA level was from 0.8 to 15.1 mg/dL. About 5.9% of participants had gout. All factors had significant statistical differences except education, details are shown in Table 1.

**Table 1.**
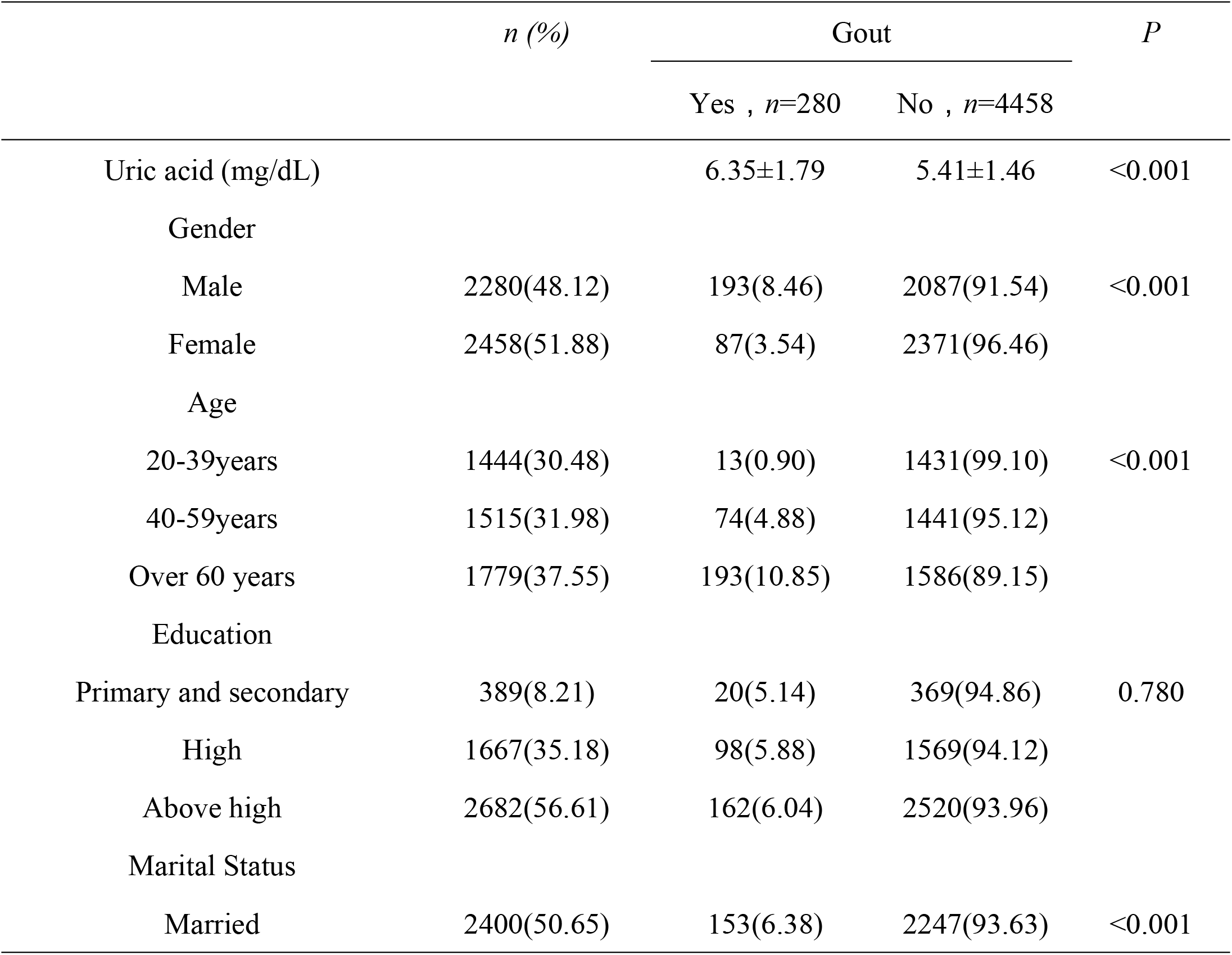
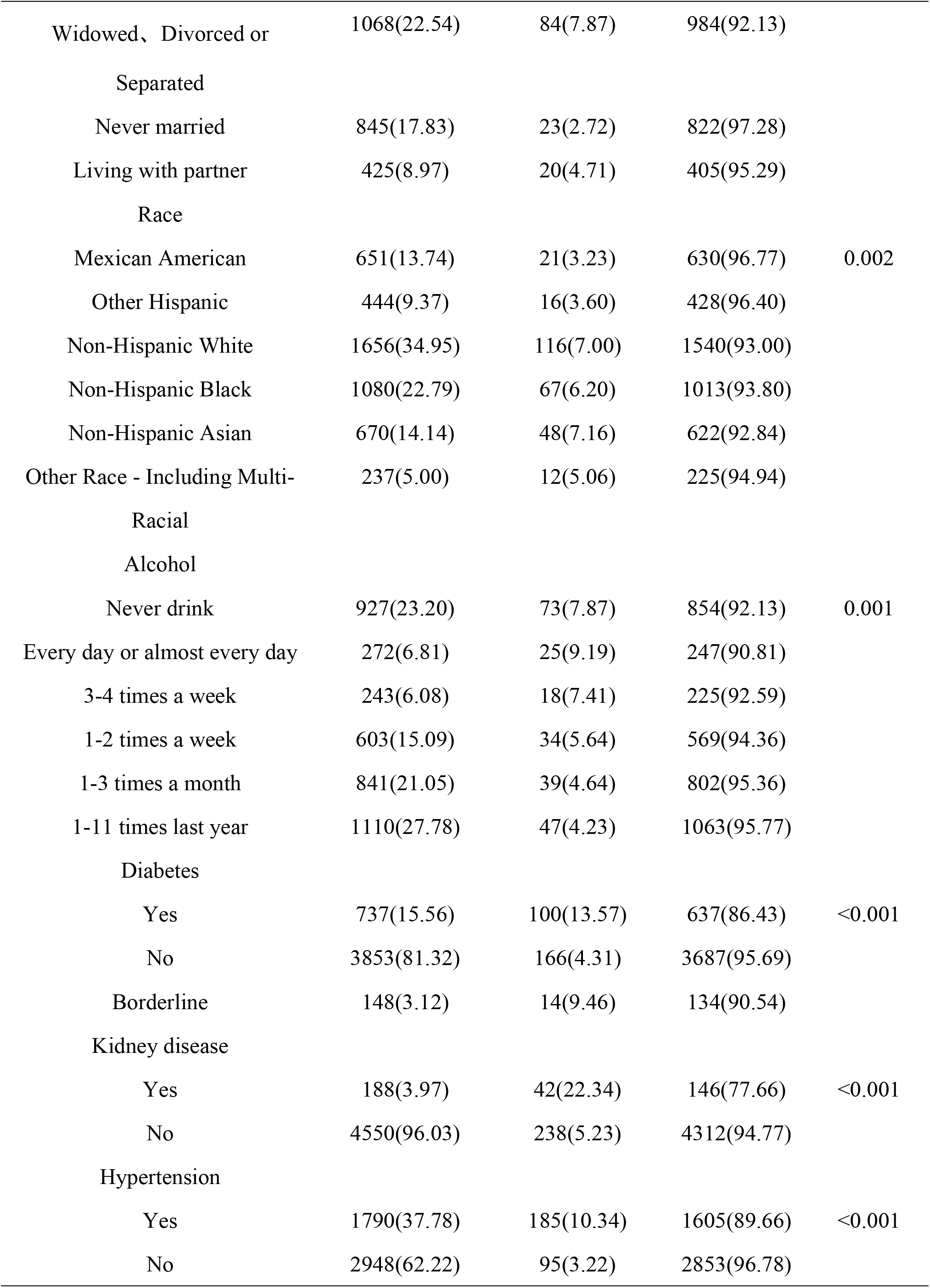
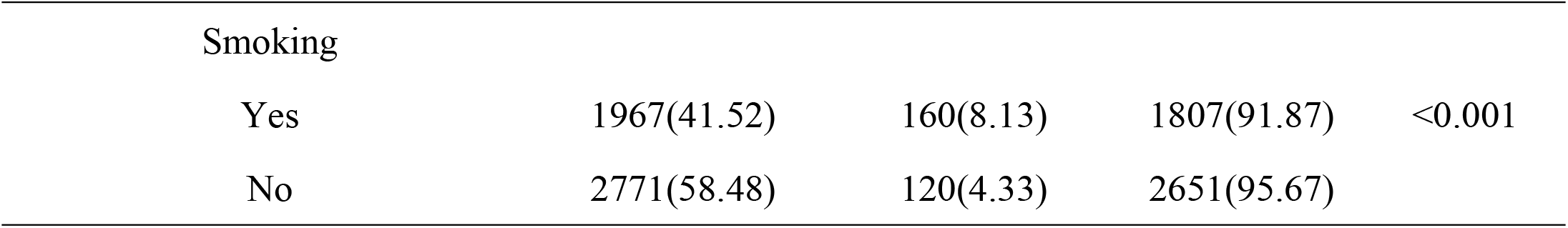
characteristics of the participants in NHANES according to gout.

|  | *n* (%) | Gout |  | *P* |
| --- | --- | --- | --- | --- |
|  |  | Yes , *n*=280 | No , *n*=4458 |  |
| Uric acid (mg/dL) |  | 6.35±1.79 | 5.41±1.46 | <0.001 |
| Gender |  |  |  |  |
| Male | 2280(48.12) | 193(8.46) | 2087(91.54) | <0.001 |
| Female | 2458(51.88) | 87(3.54) | 2371(96.46) |  |
| Age |  |  |  |  |
| 20-39years | 1444(30.48) | 13(0.90) | 1431(99.10) | <0.001 |
| 40-59years | 1515(31.98) | 74(4.88) | 1441(95.12) |  |
| Over 60 years | 1779(37.55) | 193(10.85) | 1586(89.15) |  |
| Education |  |  |  |  |
| Primary and secondary | 389(8.21) | 20(5.14) | 369(94.86) | 0.780 |
| High | 1667(35.18) | 98(5.88) | 1569(94.12) |  |
| Above high | 2682(56.61) | 162(6.04) | 2520(93.96) |  |
| Marital Status |  |  |  |  |
| Married | 2400(50.65) | 153(6.38) | 2247(93.63) | <0.001 |
| Widowed、 Divorced or<br>Separated | 1068(22.54) | 84(7.87) | 984(92.13) |  |
| Never married | 845(17.83) | 23(2.72) | 822(97.28) |  |
| Living with partner | 425(8.97) | 20(4.71) | 405(95.29) |  |
| Race |  |  |  |  |
| Mexican American | 651(13.74) | 21(3.23) | 630(96.77) | 0.002 |
| Other Hispanic | 444(9.37) | 16(3.60) | 428(96.40) |  |
| Non-Hispanic White | 1656(34.95) | 116(7.00) | 1540(93.00) |  |
| Non-Hispanic Black | 1080(22.79) | 67(6.20) | 1013(93.80) |  |
| Non-Hispanic Asian | 670(14.14) | 48(7.16) | 622(92.84) |  |
| Other Race - Including Multi-Racial | 237(5.00) | 12(5.06) | 225(94.94) |  |
| Alcohol |  |  |  |  |
| Never drink | 927(23.20) | 73(7.87) | 854(92.13) | 0.001 |
| Every day or almost every day | 272(6.81) | 25(9.19) | 247(90.81) |  |
| 3-4 times a week | 243(6.08) | 18(7.41) | 225(92.59) |  |
| 1-2 times a week | 603(15.09) | 34(5.64) | 569(94.36) |  |
| 1-3 times a month | 841(21.05) | 39(4.64) | 802(95.36) |  |
| 1-11 times last year | 1110(27.78) | 47(4.23) | 1063(95.77) |  |
| Diabetes |  |  |  |  |
| Yes | 737(15.56) | 100(13.57) | 637(86.43) | <0.001 |
| No | 3853(81.32) | 166(4.31) | 3687(95.69) |  |
| Borderline | 148(3.12) | 14(9.46) | 134(90.54) |  |
| Kidney disease |  |  |  |  |
| Yes | 188(3.97) | 42(22.34) | 146(77.66) | <0.001 |
| No | 4550(96.03) | 238(5.23) | 4312(94.77) |  |
| Hypertension |  |  |  |  |
| Yes | 1790(37.78) | 185(10.34) | 1605(89.66) | <0.001 |
| No | 2948(62.22) | 95(3.22) | 2853(96.78) |  |
| Smoking |  |  |  |  |
| Yes | 1967(41.52) | 160(8.13) | 1807(91.87) | <0.001 |
| No | 2771(58.48) | 120(4.33) | 2651(95.67) |  |

### Model 1: Gout and serum uric acid(univariable)

Model 1 used gout (Yes/No) as a binary response. The results of non-linear parameters of GAM were presented in Table 2. A smoothing spline function of SUA level as a univariable predictor was shown in Fig 2(*EDF*= 3.881, *p*<0.001). The prevalence of gout increased with the increase of SUA level at the SUA level of 5-10 mg/dL. When the SUA level was extreme, the trend was relatively flat.

**Table 2.** the smooth function of gout measured by the GAM (model1)

| Smooth terms | EDF | Ref.df | $c^2$ | $P$ |
| --- | --- | --- | --- | --- |
| Reference (Yes) |  |  |  |  |
| Gout (No) | 3.881 | 4.875 | 123.4 | <0.001 |

**Fig 2.**
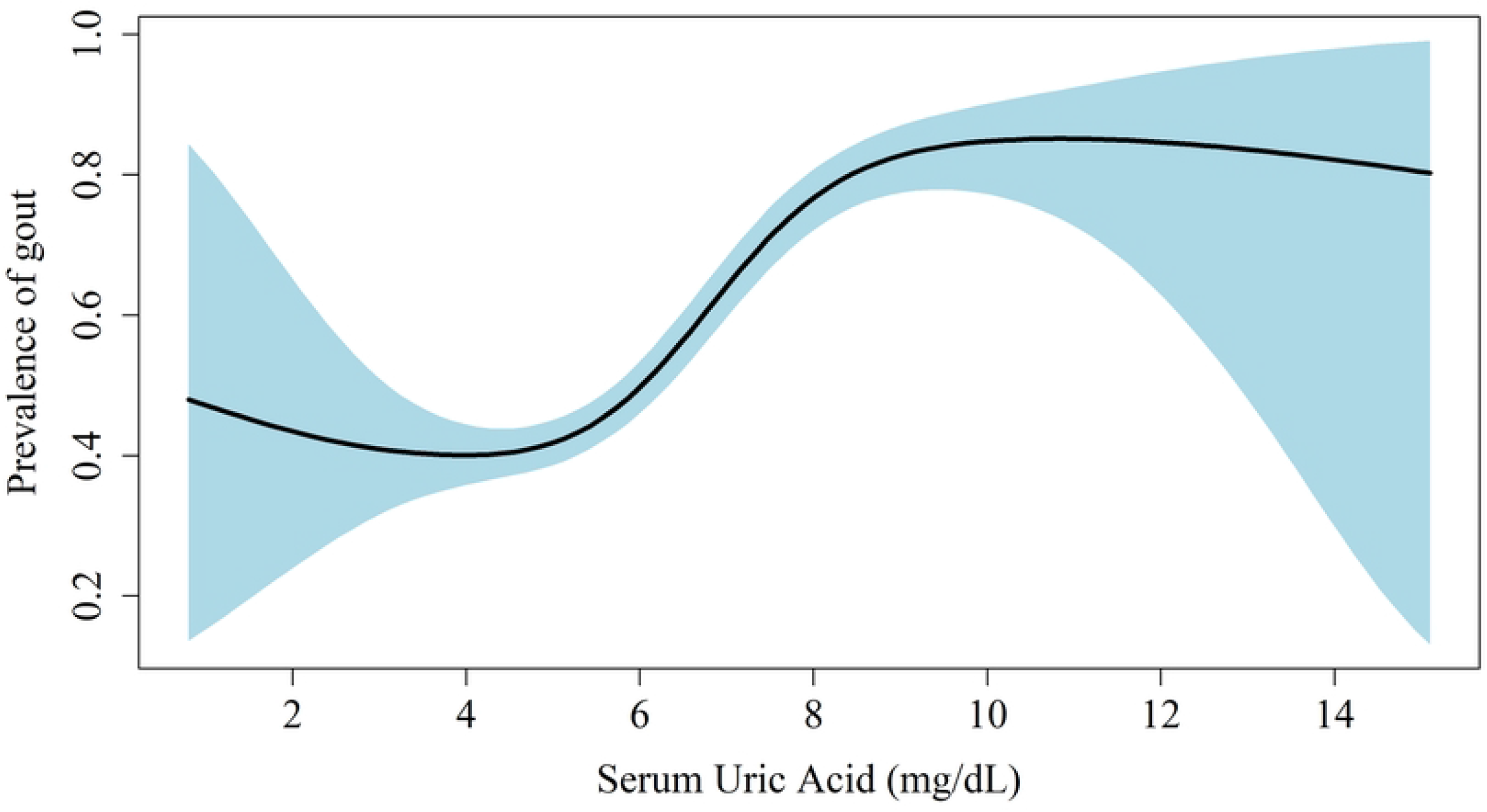
Figure of estimated smoothing spline function of prevalence of gout (The reference group is participants without gout) with 95% confidence band for the univariable generalized additive model 1.

### Model 2: Gout and serum uric acid(multivariable)

Model 2, which was a multivariable version of model 1 but further adjusted for covariates was shown in Fig 3.A non-linear shape was found between SUA level and gout (*EDF*=3.795, *p*<0.001), and the predicted smooth association of SUA level and gout including the 95% confidence intervals, was shown in Fig 3. The figure showed a U-shape association of SUA level and the prevalence of gout in multivariable GAM models. The lowest prevalence observed was SUA level of 5-6mg/dL. SUA level had the strongest effect on gout at the SUA level which was lower than 5 mg/dL or at 6-11 mg/dL. While SUA level exceed 11 mg/dL, the prevalence of gout began to decline slowly. The results of non-linear parameters of GAM were presented in Table 3, the parameter section results were detailed in S1 Table.

**Fig 3.**
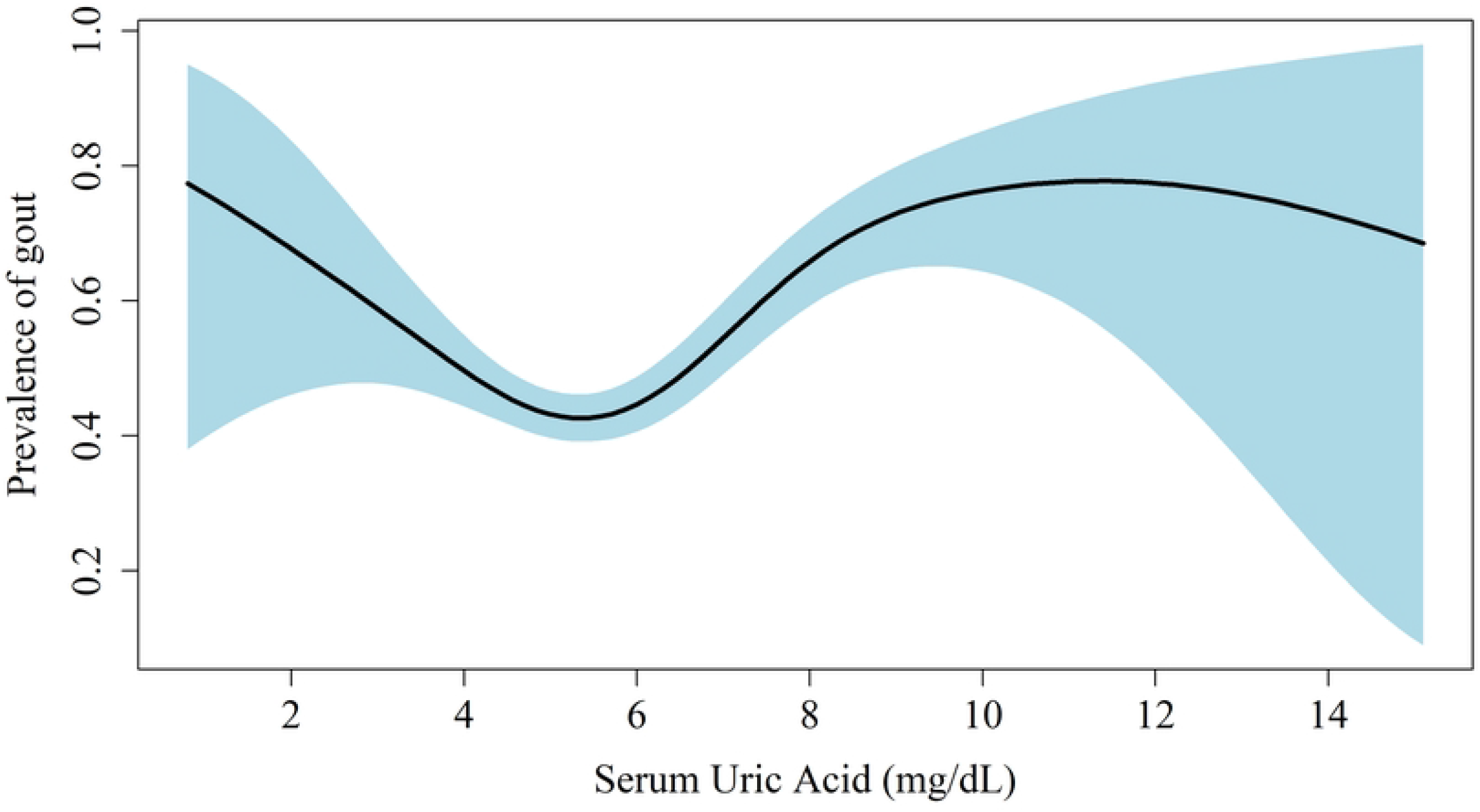
Figure of estimated smoothing spline function of prevalence of gout (The reference group is participants without gout) with 95% confidence band for the multivariable generalized additive model 2 adjusted for gender, age, marital status, education, race, smoking, alcohol, diabetes, kidney disease and hypertension.

**Table 3.** the smooth function of gout measured by the GAM (model2)

| Smooth terms | EDF | Ref.df | $c^2$ | $P$ |
| --- | --- | --- | --- | --- |
| Reference (Yes) |  |  |  |  |
| Gout (No) | 3.795 | 4.789 | 37.67 | <0.001 |

### Model 3: Gout and serum uric acid (Group by gender)

Furthermore, the effect of SUA level on gout in male and female was evaluated in model 3. The significant result indicates a non-linear shape between SUA level and gout in male (*EDF*=1.873, *p*<0.001), which was similar to the trend of entire population. However, the non-linear shape between SUA level and gout was not identified in female (*EDF*=1.001, *p*=0.087). The Fig 4 showed the smoothing shapes are different in male and female, which a significant U-shape association of SUA level and the gout in male and a linear in female. The lowest risk is observed in those SUA level 5∼6mg/dL in male and low or high SUA level will increase the risk of gout. The results of non-linear parameters of GAM were presented in Table 4, the parameter section results were detailed in S2.1 and S2.2 Table.

**Fig 4.**
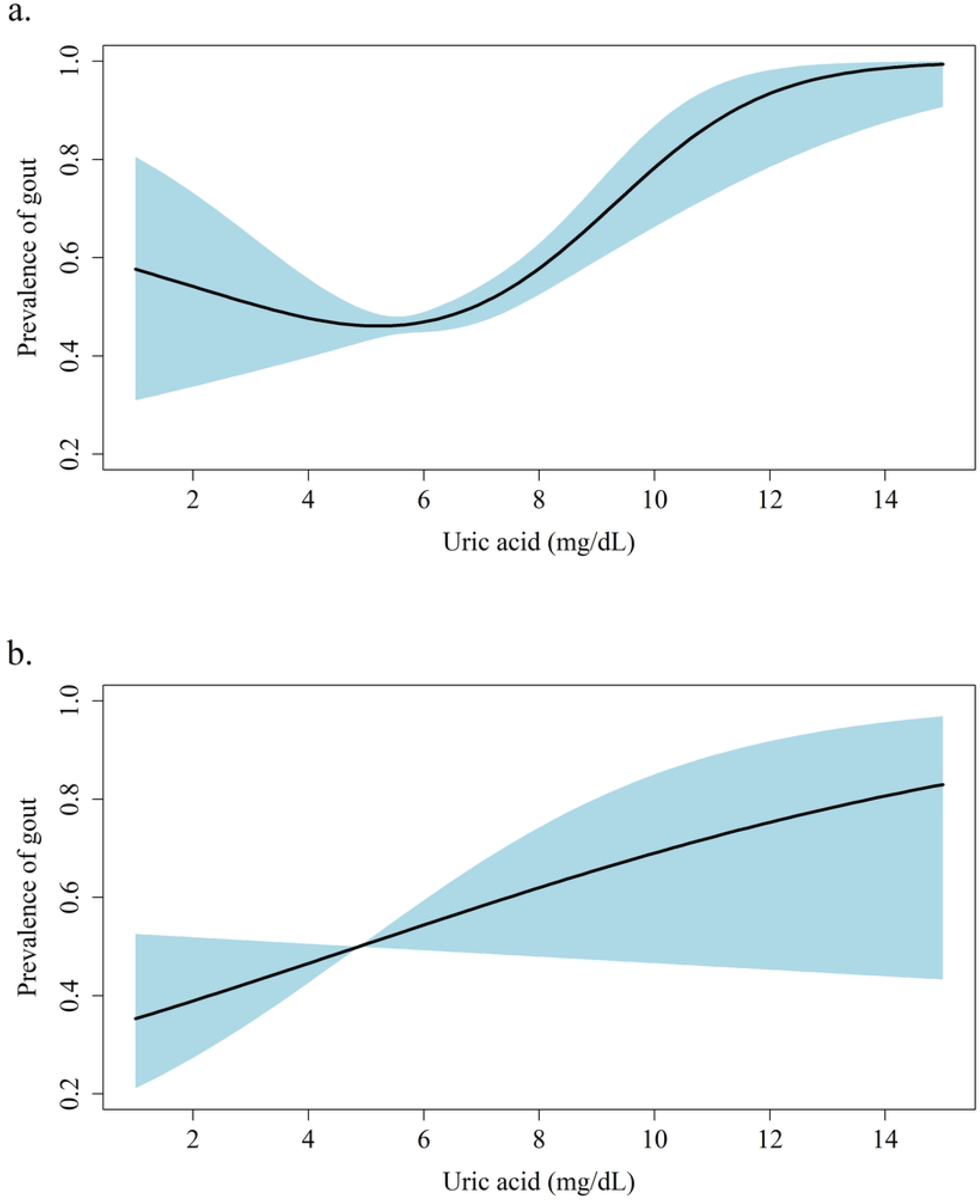
Figures of estimated smoothing spline function of prevalence of gout level (Group by gender, and the reference group is participants without gout) with 95% confidence band for the multivariable generalized additive model 3. And a for male, b for female.

**Table 4.** Association of SUA level and gout measured by the GAM(Group by gender)(model3)

| Smooth terms | EDF | Ref.df | $c^2$ | $P$ |
| --- | --- | --- | --- | --- |
| Reference (Yes) |  |  |  |  |
| Spline (Uric Acid): Male | 1.873 | 1.984 | 16.45 | <0.001 |
| Spline (Uric Acid): Female | 1.001 | 1.002 | 2.931 | 0.087 |

## Discussion

As the final product of purine metabolism, SUA has many effects on the human body. Previous studies found that increased SUA levels were associated with gout, diabetes[20], hypertension[21], obesity[22], and cancer[23, 24]. However, few studies have assessed the relationship between across-full range of SUA level and gout. This study constructs a generalized additivity model to explore the potential relationship between SUA level and gout based on US adults.

In our study, the *EDF* of univariate and multivariate models adjusted for age, sex, education and other important confounding factors were all greater than 3, indicating a non-linear relationship between SUA level and gout and a U-shape association was demonstrated in the figure. The results of our study showed a high prevalence of gout with high SUA level in US adults, but the prevalence of gout begins to decline slowly when SUA level is above 11 mg/dL. Males with low SUA level also have a high prevalence of gout. The association between SUA level and prevalence of gout support a U-shape in males, while females show a linear shape.

A previous study already confirmed that SUA level higher than 6mg/dL can increase the risk of gout, which is in agreement with our result[25]. Secondly, the prevalence of gout decreased when SUA level was higher than 11 mg/dL. We speculated that the high level of SUA in this part of the population may be caused by other diseases or drugs[26]. Furthermore, low SUA level was also associated with gout. A sudden decrease in SUA may also trigger a gout attack by dissolution of monosodium urate (MSU) from tophi[26]. In addition, hypouricemia has been shown to increase the risk of worsening kidney function. In the CARES trial, all-cause and cardiovascular mortality rates were higher in the febuxostat group than in the allopurinol group, despite lower SUA levels in gout patients taking febuxostat[27]. Some previous studies also showed that SUA was u-shape associated with all-cause mortality[28, 29] and nephropathy, and SUA reduction treatment was associated with higher mortality[30].One study showed that SUA level between 5-6mg/dL is reasonable[31], which is consistent with our finding. Ames et al hypothesized that SUA may act as a protective factor against aging and cancer caused by oxidants and free radicals due to its antioxidant function[32]. Therefore, SUA should be maintained in a certain range, above or below this range will increase the probability of developing gout. The different trends between males and females are because estrogen has a urination effect on females, which can reduce the risk of gout. However, with elevated SUA level after menopause, the proportion of older female with gout may increase and gender differences decrease[17, 33].

Studies have shown that the occurrence of gout is related to kidney disease[9, 34, 35]. SUA level in vivo is closely related to renal function. About two-thirds of uric acid is excreted in urine[36], and uric acid is filtered by glomerulus and reabsorbed by proximal tubules, with a normal excretion rate of 10%[37]. A prospective cohort study showed a U-shape association between SUA and loss of renal function in healthy subjects[38]. Therefore, the relationship between SUA and kidney disease may be a potential cause for the existence of this U-shape association that was found in our study, but further studies are needed to prove this. Because gout is considered a disease that primarily affects male, it may be underrecognized in female even after menopause. There were clear differences in risk between male and female.

There are also limitations in our research. Firstly, our research is an observational study, which cannot infer a causal relationship between SUA level and gout. Secondly, many variables in our study are self-reported. Despite these limitations, our study also has advantages. Firstly, this study used a smoothing curve to explore the non-linear trend between SUA level and gout. Secondly, we assessed the risk of gout at all SUA level in the ordinary population, taking into account both low SUA level and high SUA level. Finally, we assessed the trend of prevalence of gout with SUA level in both males and females.

## Conclusion

In our study, there is a U-shape between SUA level and gout. High SUA level or low SUA level are both risk factors for gout. Further researches are needed to trace people with low SUA level and clarify the pathophysiological mechanisms between low SUA level and gout to reduce the likelihood of gout occurring.

## Data Availability

Data are available in a public, open access repository. The datasets for this study can be found in https://wwwn.cdc.gov/Nchs/Nhanes/2017-2018/index.htm.

https://www.cdc.gov/Nchs/Nhanes/2017-2018/index.htm

## Author Contributions

YXM and STL were responsible for data extraction, data analyses, model construction and the manuscript writing. LMJ selected the topic, directs the writing, improves the revision. ZZX provided specific knowledge, carried out the literature research. ZHB, ZH, LYR coordinated and managed the operation of the project and the implementation of the project. All authors read and approved the final manuscript.

## References

1. Safiri S, Kolahi A-A, Cross M, Carson-Chahhoud K, Hoy D, Almasi-Hashiani A, et al. Prevalence, Incidence, and Years Lived With Disability Due to Gout and Its Attributable Risk Factors for 195 Countries and Territories 1990-2017: A Systematic Analysis of the Global Burden of Disease Study 2017. Arthritis Rheumatol. 2020;72(11):1916–27. doi: 10.1002/art.41404. PubMed PMID: 32755051.

2. Disveld IJM, Zoakman S, Jansen TLTA, Rongen GA, Kienhorst LBE, Janssens HJEM, et al. Crystal-proven gout patients have an increased mortality due to cardiovascular diseases, cancer, and infectious diseases especially when having tophi and/or high serum uric acid levels: a prospective cohort study. Clin Rheumatol. 2019;38(5):1385–91. doi: 10.1007/s10067-019-04520-6. PubMed PMID: 30929152.

3. Bardin T, Richette P. Impact of comorbidities on gout and hyperuricaemia: an update on prevalence and treatment options. BMC Med. 2017;15(1):123. doi: 10.1186/s12916-017-0890-9. PubMed PMID: 28669352.

4. Roddy E, Zhang W, Doherty M. Is gout associated with reduced quality of life? A case-control study. Rheumatology (Oxford). 2007;46(9):1441-4. PubMed PMID: 17586863.

5. Edwards NL, Sundy JS, Forsythe A, Blume S, Pan F, Becker MA. Work productivity loss due to flares in patients with chronic gout refractory to conventional therapy. J Med Econ. 2011;14(1):10–5. doi: 10.3111/13696998.2010.540874. PubMed PMID: 21138339.

6. Kleinman NL, Brook RA, Patel PA, Melkonian AK, Brizee TJ, Smeeding JE, et al. The impact of gout on work absence and productivity. Value Health. 2007;10(4):231-7. PubMed PMID: 17645677.

7. Khanna D, Ahmed M, Yontz D, Ginsburg SS, Park GS, Leonard A, et al. The disutility of chronic gout. Qual Life Res. 2008;17(5):815–22. doi: 10.1007/s11136-008-9355-0. PubMed PMID: 18500578.

8. Singh JA. Racial and gender disparities among patients with gout. Curr Rheumatol Rep. 2013;15(2):307. doi: 10.1007/s11926-012-0307-x. PubMed PMID: 23315156.

9. Kuo C-F, Grainge MJ, Zhang W, Doherty M. Global epidemiology of gout: prevalence, incidence and risk factors. Nat Rev Rheumatol. 2015;11(11):649–62. doi: 10.1038/nrrheum.2015.91. PubMed PMID: 26150127.

10. Richette P, Bardin T. Gout. Lancet. 2010;375(9711):318–28. doi: 10.1016/S0140-6736(09)60883-7. PubMed PMID: 19692116.

11. Dalbeth N, Gosling AL, Gaffo A, Abhishek A. Gout. Lancet. 2021;397(10287):1843–55. doi: 10.1016/S0140-6736(21)00569-9. PubMed PMID: 33798500.

12. Hainer BL, Matheson E, Wilkes RT. Diagnosis, treatment, and prevention of gout. Am Fam Physician. 2014;90(12):831-6. PubMed PMID: 25591183.

13. Perez-Ruiz F. Treating to target: a strategy to cure gout. Rheumatology (Oxford). 2009;48 Suppl 2. doi: 10.1093/rheumatology/kep087. PubMed PMID: 19447780.

14. Koto R, Nakajima A, Horiuchi H, Yamanaka H. Serum uric acid control for prevention of gout flare in patients with asymptomatic hyperuricaemia: a retrospective cohort study of health insurance claims and medical check-up data in Japan. Ann Rheum Dis. 2021;80(11):1483–90. doi: 10.1136/annrheumdis-2021-220439. PubMed PMID: 34158371.

15. Keenan T, Zhao W, Rasheed A, Ho WK, Malik R, Felix JF, et al. Causal Assessment of Serum Urate Levels in Cardiometabolic Diseases Through a Mendelian Randomization Study. J Am Coll Cardiol. 2016;67(4):407–16. doi: 10.1016/j.jacc.2015.10.086. PubMed PMID: 26821629.

16. Ruoff G, Edwards NL. Overview of Serum Uric Acid Treatment Targets in Gout: Why Less Than 6 mg/dL? Postgrad Med. 2016;128(7):706–15. doi: 10.1080/00325481.2016.1221732. PubMed PMID: 27558643.

17. Saag KG, Choi H. Epidemiology, risk factors, and lifestyle modifications for gout. Arthritis Res Ther. 2006;8 Suppl 1:S2. PubMed PMID: 16820041.

18. Wallace KL, Riedel AA, Joseph-Ridge N, Wortmann R. Increasing prevalence of gout and hyperuricemia over 10 years among older adults in a managed care population. J Rheumatol. 2004;31(8):1582-7. PubMed PMID: 15290739.

19. D’Silva KM, Yokose C, Lu N, McCormick N, Lee H, Zhang Y, et al. Hypouricemia and Mortality Risk in the US General Population. Arthritis Care Res (Hoboken). 2021;73(8):1171–9. doi: 10.1002/acr.24476. PubMed PMID: 33026684.

20. van der Schaft N, Brahimaj A, Wen K-X, Franco OH, Dehghan A. The association between serum uric acid and the incidence of prediabetes and type 2 diabetes mellitus: The Rotterdam Study. PLoS One. 2017;12(6):e0179482. doi: 10.1371/journal.pone.0179482. PubMed PMID: 28632742.

21. Zheng R, Yang T, Chen Q, Chen C, Mao Y. Serum Uric Acid Concentrations Can Predict Hypertension: A Longitudinal Population-Based Epidemiological Study. Horm Metab Res. 2017;49(11):873–9. doi: 10.1055/s-0043-119129. PubMed PMID: 28922678.

22. Zheng R, Chen C, Yang T, Chen Q, Lu R, Mao Y. Serum Uric Acid Levels and the Risk of Obesity: a Longitudinal Population-Based Epidemiological Study. Clin Lab. 2017;63(10):1581–7. doi: 10.7754/Clin.Lab.2017.170311. PubMed PMID: 29035437.

23. Yue C-F, Feng P-N, Yao Z-R, Yu X-G, Lin W-B, Qian Y-M, et al. High serum uric acid concentration predicts poor survival in patients with breast cancer. Clin Chim Acta. 2017;473:160–5. doi: 10.1016/j.cca.2017.08.027. PubMed PMID: 28844462.

24. Shin H-S, Lee H-R, Lee D-C, Shim J-Y, Cho K-H, Suh S-Y. Uric acid as a prognostic factor for survival time: a prospective cohort study of terminally ill cancer patients. J Pain Symptom Manage. 2006;31(6):493-501. PubMed PMID: 16793489.

25. Perez-Ruiz F, Dalbeth N, Bardin T. A review of uric acid, crystal deposition disease, and gout. Adv Ther. 2015;32(1):31–41. doi: 10.1007/s12325-014-0175-z. PubMed PMID: 25533440.

26. Zhang W-Z. Why Does Hyperuricemia Not Necessarily Induce Gout? Biomolecules. 2021;11(2). doi: 10.3390/biom11020280. PubMed PMID: 33672821.

27. White WB, Saag KG, Becker MA, Borer JS, Gorelick PB, Whelton A, et al. Cardiovascular Safety of Febuxostat or Allopurinol in Patients with Gout. N Engl J Med. 2018;378(13):1200–10. doi: 10.1056/NEJMoa1710895. PubMed PMID: 29527974.

28. Hu L, Hu G, Xu BP, Zhu L, Zhou W, Wang T, et al. U-Shaped Association of Serum Uric Acid With All-Cause and Cause-Specific Mortality in US Adults: A Cohort Study. J Clin Endocrinol Metab. 2020;105(1). doi: 10.1210/clinem/dgz068. PubMed PMID: 31650159.

29. Stotz M, Szkandera J, Seidel J, Stojakovic T, Samonigg H, Reitz D, et al. Evaluation of uric acid as a prognostic blood-based marker in a large cohort of pancreatic cancer patients. PLoS One. 2014;9(8):e104730. doi: 10.1371/journal.pone.0104730. PubMed PMID: 25133546.

30. Odden MC, Amadu A-R, Smit E, Lo L, Peralta CA. Uric acid levels, kidney function, and cardiovascular mortality in US adults: National Health and Nutrition Examination Survey (NHANES) 1988-1994 and 1999-2002. Am J Kidney Dis. 2014;64(4):550–7. doi: 10.1053/j.ajkd.2014.04.024. PubMed PMID: 24906981.

31. Bellomo G, Selvi A. Uric Acid: The Lower the Better? Contrib Nephrol. 2018;192:69–76. doi: 10.1159/000484280. PubMed PMID: 29393097.

32. Ames BN, Cathcart R, Schwiers E, Hochstein P. Uric acid provides an antioxidant defense in humans against oxidant-and radical-caused aging and cancer: a hypothesis. Proc Natl Acad Sci U S A. 1981;78(11):6858-62. PubMed PMID: 6947260.

33. Doherty M. New insights into the epidemiology of gout. Rheumatology (Oxford). 2009;48 Suppl 2:ii2–ii8. doi: 10.1093/rheumatology/kep086. PubMed PMID: 19447779.

34. Feig DI, Kang D-H, Johnson RJ. Uric acid and cardiovascular risk. N Engl J Med. 2008;359(17):1811–21. doi: 10.1056/NEJMra0800885. PubMed PMID: 18946066.

35. Dehlin M, Jacobsson L, Roddy E. Global epidemiology of gout: prevalence, incidence, treatment patterns and risk factors. Nat Rev Rheumatol. 2020;16(7):380–90. doi: 10.1038/s41584-020-0441-1. PubMed PMID: 32541923.

36. Bobulescu IA, Moe OW. Renal transport of uric acid: evolving concepts and uncertainties. Adv Chronic Kidney Dis. 2012;19(6):358–71. doi: 10.1053/j.ackd.2012.07.009. PubMed PMID: 23089270.

37. Fathallah-Shaykh SA, Cramer MT. Uric acid and the kidney. Pediatr Nephrol. 2014;29(6). doi: 10.1007/s00467-013-2549-x. PubMed PMID: 23824181.

38. Kanda E, Muneyuki T, Kanno Y, Suwa K, Nakajima K. Uric acid level has a U-shaped association with loss of kidney function in healthy people: a prospective cohort study. PLoS One. 2015;10(2):e0118031. doi: 10.1371/journal.pone.0118031. PubMed PMID: 25658588.

